# Systematic review and meta-analysis reveal positive therapeutic effects of music in brain damage rehabilitation

**DOI:** 10.1101/2025.07.29.25332404

**Authors:** Laura Navarro, Nour El Zahraa Mallah, Jacobo Pardo-Seco, Alberto Gómez-Carballa, Sara Pischedda, Wiktor Nowak, Emma Segura, Antoni Rodriguez-Fornells, Federico Martinón-Torres, Antonio Salas

## Abstract

**Importance:** Brain damage often results in impairments across motor, cognitive, communicative, and psychosocial domains. Identifying effective, accessible rehabilitation strategies is critical for improving recovery and quality of life.

**Objective:** To systematically review and synthesize the evidence on the therapeutic effects of music-based interventions in individuals with brain damage, with a focus on functional outcomes and potential neurobiological mechanisms.

**Evidence Review:** This systematic review and meta-analysis evaluated studies examining music-based interventions in populations with brain damage. The analysis included a total of 70 randomized and non-randomized controlled trials, focusing on motor, cognitive, communicative, emotional, behavioral, and social outcomes. Studies were assessed for methodological rigor, and data were synthesized both qualitatively and quantitatively.

**Findings:** Music-based interventions consistently supported motor recovery, particularly in gait and upper-limb function, likely through mechanisms such as rhythmic entrainment and auditory-motor coupling. Cognitive outcomes, including memory, attention, and executive function, showed robust improvements, often linked to music-induced neuroplasticity in frontal and temporal brain regions. Communicative and psychosocial outcomes were less frequently studied but demonstrated benefits in language production, mood regulation, and emotional well-being. Behavioral improvements, including self-care and coping, showed positive trends, though many did not reach statistical significance. Social functioning was indirectly enhanced through improved emotional expression. Despite brain damage-related impairments such as amusia, many patients retained musical memory and responsiveness, with prior musical training possibly conferring neuroprotective effects. Methodological limitations, including small sample sizes and heterogeneity in intervention design, restrict generalizability.

**Conclusions and Relevance:** Music-based interventions appear to be a promising, multimodal intervention for individuals with brain damage, yielding improvements across multiple functional domains. While evidence supports its clinical utility, future research should focus on larger, more standardized trials to clarify mechanisms, optimize protocols, and confirm long-term benefits. Integration of music-based interventions into neurorehabilitation programs may enhance recovery through targeted modulation of neurobiological pathways.

## 1. Introduction

Music is an integral part of daily life across all ages and cultures, serving as one of the most universal forms of expression and communication ^1^. While cultural and educational perspectives highlight the significance of musical stimuli for human development and well-being ^2^, scientific exploration of music’s biological effects remains relatively limited. Music is an extremely complex auditory stimulus, with dynamic changes over time in various acoustic sound features, including timbre, intensity, frequency, and tempo, representing a complex version of the acoustic environment. Over the past two decades, cognitive sciences and neurosciences have shown increasing interest in music as a highly complex and versatile stimulus, offering valuable insights into brain functions ^3,4^. Relevant studies have shown the plasticity of the human auditory cortex related to musical training ^5^, and how musical processing involves a widespread network of brain structures in addition to the auditory cortex, including the cerebellum, planum temporale, parietal lobe, insula, limbic circuit, ventral striatum (nucleus accumbens), ventral tegmental area, premotor cortex, anterior superior-temporal gyrus, and frontal lobe, among others ^6^. In genetics, research has identified specific regions and genes linked to musical traits ^7^ and explored the heritability of musical characteristics ^8^.

Stroke has been identified as the leading contributor to neurological diseases in a recent report, with its incidence increasing by 86.1% from 1990 to 2021 ^9^. Cerebral palsy, as both a health and social issue, demands innovative approaches to explore the potential of non-pharmacological interventions in improving patients’ quality of life (QoL). In this context, music has gained increasing attention for its potential use for clinical interventions, particularly in neurodegenerative conditions ^10^ and for addressing brain damage (BD) resulting from stroke or other acquired brain injuries (ABI). Previous reviews, mostly systematic or narrative (qualitative), with only a few including meta-analyses on specific outcomes, have focused on the rehabilitative effects of music, particularly examining specific outcomes such as the effectiveness of music therapy in traumatic brain injury (TBI) ^11^, the effects of choral singing on BD ^12^, the benefits of active music-based interventions (MI) in upper-limb rehabilitation ^13^, or the impact of previous musical training following BD ^14^. A key systematic review ^15^ comprehensively analyzed the effects of MI on BD, while other reviews have focused on specific outcomes, such as gait improvement ^16^ or language recovery^17^.

Our study represents the first comprehensive and integrative systematic review and meta-analysis on the multifaceted impact of MI in BD rehabilitation. By bridging motor, cognitive, and well-being domains, we synthesize a wide range of research through both qualitative analysis and quantitative meta-analytic methods. This dual approach deepens interpretive insight and offers robust statistical evidence, shedding new light on the complex and often underestimated interplay between music and neurological recovery.

## 2. Methods

A systematic review was conducted using PubMed (initial search completed on March 24, 2023). Search terms included: “brain injur”, “brain deformation”, “cerebral damage”, and “music”, using the [tiab] field to restrict results to title and abstract. This search yielded 247 records. After applying filters for human studies in English, published from 2000 onward, and excluding reviews or meta-analyses, 92 articles remained. Following detailed screening, 23 studies met the inclusion criteria. An additional 34 were added *via* reference mining. To ensure completeness, a second search on June 11, 2025, expanded the scope to PubMed, Embase, Scopus, Cochrane Library, Web of Science, and ClinicalTrials.gov. The same search strategy was applied, limited to 2023–2025 publications. Of 311 records, 153 unique studies remained after removing 158 duplicates (*via* Rayyan). Following manual screening and full-text review, 9 studies were retrieved, and 4 more were included after expert consultation, yielding a final total of 70 publications. The review followed PRISMA guidelines (https://www.bmj.com/content/372/bmj.n71), **Figure 1**.

**Figure 1.**
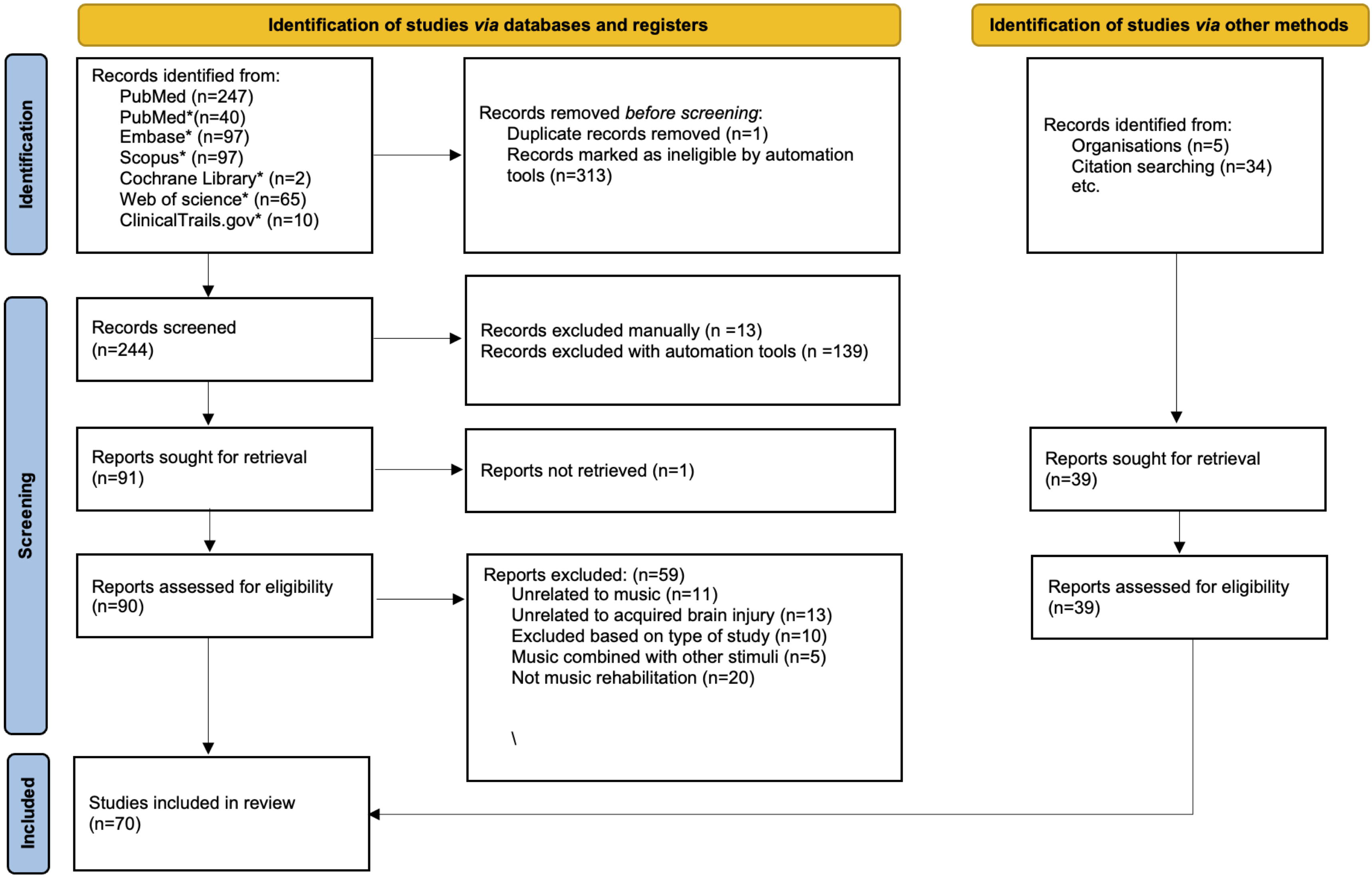
PRISMA flow diagram of study selection. This diagram outlines the systematic identification, screening, eligibility assessment, and final inclusion of studies in accordance with PRISMA guidelines. The process includes records identified *via* PubMed and citation searching, detailing the number of studies excluded at each stage and the reasons for exclusion. The flowchart follows the updated PRISMA framework proposed by Page et al. ^76^

A meta-analysis was conducted on 31 studies that provided complete data suitable for statistical synthesis. Key outcomes included general recovery, motor and cognitive function, communication, emotional well-being, and QoL. Data were independently extracted by two reviewers and included sample sizes, means, standard deviations, intervention durations, and outcome measures. Studies were excluded if they lacked a control group, did not isolate the specific effects of MI, or focused solely on acute physiological outcomes. Given the substantial heterogeneity observed across studies (I^2^>50% in several domains), a random-effects model was employed using the DerSimonian and Laird method ^18^, which accounts for variability across diverse populations, interventions, and study designs. Heterogeneity was assessed using the I^2^, *τ*^2^, and Cochran’s Q. Where significant heterogeneity was detected, subgroup analyses were conducted, and subgroup differences were evaluated using *χ*^2^ tests. Missing standard deviations were imputed using r=0.8, in line with Cochrane recommendations ^19,20^. Forest plots were generated to illustrate individual study outcomes and pooled effect sizes. To ensure interpretability, effect directions were harmonized across measurement scales, and standardized metrics were applied when necessary. Some studies were excluded from specific subgroup analyses due to incompatible outcome metrics; however, separate meta-analyses were conducted within each functional domain (e.g., UEF, gait) to maintain analytical clarity. Prediction intervals were calculated to estimate the expected range of effects in future comparable studies, incorporating both within- and between-study variance. All analyses were conducted using the META package in R ^21^. The statistical significance of pooled effects was assessed using the z-test. See **Supplementary Text 1** for further methodological details.

## 3. Results

### 3.1. Bibliographic systematic review and meta-analysis

Acquired Brain Injury (ABI), encompassing both traumatic (TBI) and non-traumatic (non-TBI) causes, presents a wide range of clinical profiles that complicate the generalization of MI. Our systematic review (**Figure 1**) identified 70 studies, including 15 focused on TBI, 39 on non-TBI, and others targeting specific diagnoses such as aphasia, agnosia, or amusia (**Table S1**). These studies featured diverse sample sizes and methodologies, including randomized controlled trials (RCTs), case reports, and neuroimaging investigations, and covered cognitive, motor, and emotional rehabilitation domains. Despite considerable heterogeneity, the majority of studies reported beneficial outcomes. A meta-analysis (n=31; **Section 3.3**) further evaluated effects by functional domain, reinforcing the rehabilitative potential of music. While evidence supports music’s capacity to promote neuroanatomical reorganization and functional recovery, broader clinical adoption is still limited by methodological inconsistencies and small sample sizes.

### 3.2. The therapeutic potential of music training in neurorehabilitation

Despite strong scientific support for music training’s therapeutic benefits, its clinical implementation remains limited. Since 2000, a total of 70 studies have examined the effects of musical stimulation in individuals with BD, consistently reporting beneficial outcomes across a range of functions (**Table S1**). However, varied study designs, participant characteristics, and intervention protocols complicate standardization and broader clinical adoption. The experimental designs included randomized controlled trials (RCTs), detailed case reports, studies utilizing neuroimaging to assess functional or structural brain changes, clinical trials, and quasi-experimental approaches. Many of these investigations assessed pre- and post-intervention effects or compared outcomes with alternative therapies or healthy control groups.

Analysis of this body of research reveals that music-based rehabilitation could improve behavioral, cognitive, and motor functions, while also promoting neuroanatomical reorganization during recovery. These therapeutic effects are categorized by outcome domains such as motor function, communication skills, cognitive performance, emotional regulation, behavior, and social engagement. To strengthen the evidence base, we conducted a meta-analysis when sufficient data were available in these studies (*n*=31), targeting specific outcome domains to provide a rigorous and quantitative synthesis of the findings. This approach enables a clearer, more systematic evaluation of music’s rehabilitative efficacy and highlights its potential as a complementary tool in neurorehabilitation.

#### 3.2.1. Motor rehabilitation benefits

Thirty-two studies demonstrate the effectiveness of MI for motor rehabilitation in individuals with BD, focusing on gait (8 studies) and UEF (26 studies). Music-Supported Therapy (MST), involving piano and percussion exercises, emerged as a powerful approach for enhancing mobility (**Table S1**).

Gait impairments are common after cerebral injury. Rhythmic Auditory Stimulation (RAS), using metronome-enhanced musical rhythms, showed significant benefits across gait parameters ^22–26^. A meta-analysis of five studies confirmed improvements in gait performance, step length, cadence, and velocity (**Figure 2**), with significant pooled mean differences (MD=5.47; 95%CI: 2.08–8.86; z=3.16; *P*<0.01), despite high heterogeneity (I^2^=96%). Subgroup analyses revealed particularly strong effects for cadence (z=8.76; *P*<0.01; I^2^=9%) and velocity (z=7.28; *P*<0.01; I^2^=26%), while balance and symmetry showed mixed or nonsignificant results. This suggests MI excels at supporting temporal aspects of walking but may need to be supplemented for postural control.

**Figure 2.**
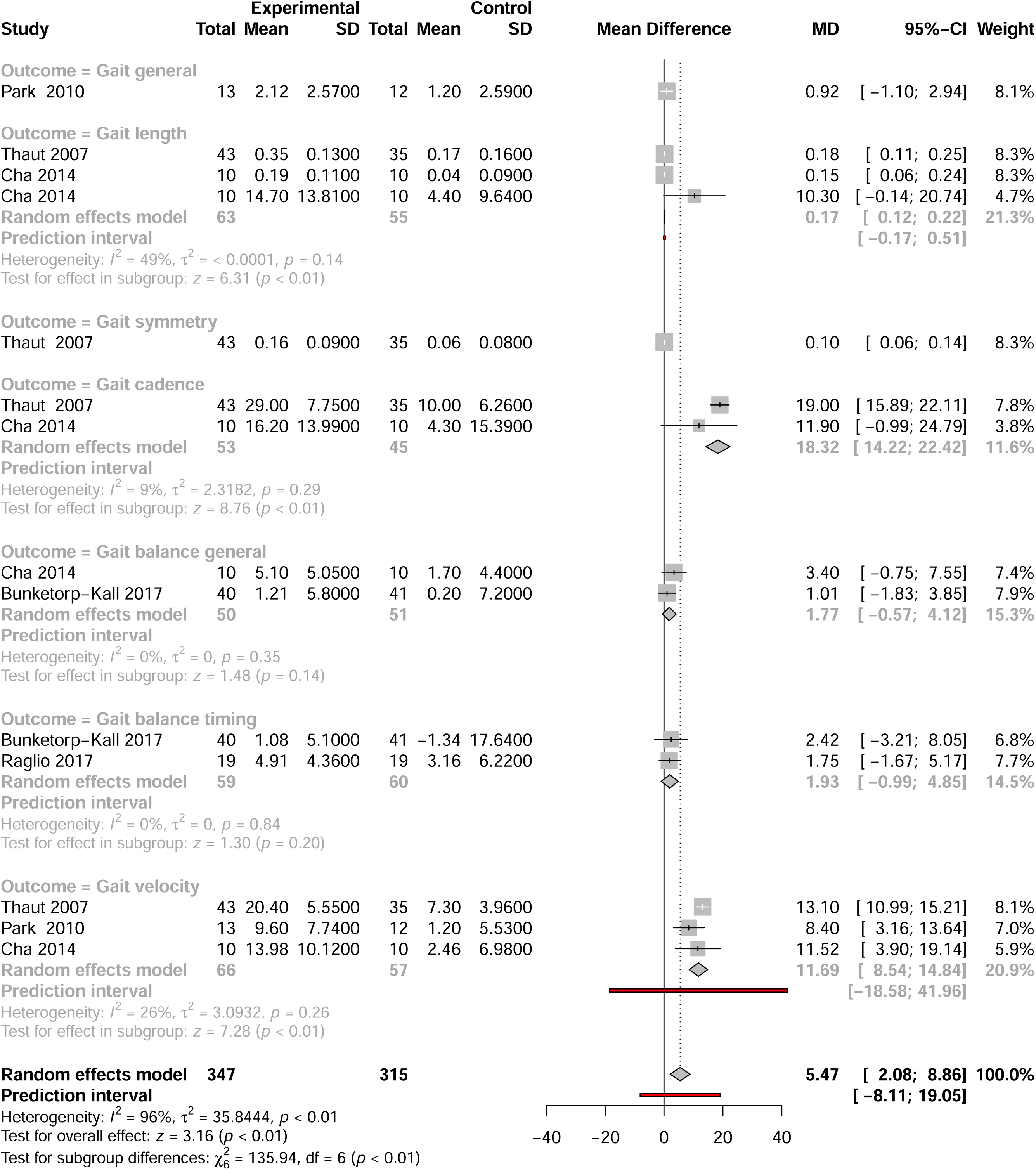
Forest plot of mean differences (MD) between experimental and control groups across studies evaluating the effects of intervention on gait-related outcomes. The plot presents individual study results with corresponding MD, 95% confidence intervals (CIs), and study weights. Summary estimates were calculated using a random-effects model. Heterogeneity statistics (I^2^, *τ*^2^, and *P*-values) are reported for each outcome, along with prediction intervals and tests for subgroup and overall effects.

Twenty-six studies support the use of MI, such as piano and drum training, for enhancing UEF through real-time sensory feedback. Reported benefits include improvements in finger speed, hand strength, shoulder flexibility, and fine motor skills ^27–30^. A meta-analysis of 12 studies (**Figure 3**) confirmed significant gains in UEF (MD=2.56; 95%CI: 1.59–3.54; z=5.18; *P*<0.01), with moderate heterogeneity (I^2^=45%) and variation across functional domains (*χ*^2^=32.28; *P*<0.01). Dexterity measures like the Box and Block Test (MD=4.51; 95%CI: 0.53–8.49; z=2.22; *P*=0.03; I^2^=60%) and Nine-Hole Pegboard Test-pegs (MD=1.15; 95%CI: 0.16– 2.14; z=2.29; *P*=0.02; I^2^=0%) showed notable improvements. Task-oriented outcomes, e.g., the ARAT (MD=5.79; 95%CI: 2.79–8.79; z=3.78; *P*<0.01) and Fugl-Meyer Assessment (MD=3.06; 95%CI: 0.27–5.85; z=2.15; *P*=0.03), demonstrated strong effects. While MI significantly enhances coordination and task performance, gains in strength and flexibility were less consistent, suggesting the need for complementary therapies.

**Figure 3.**
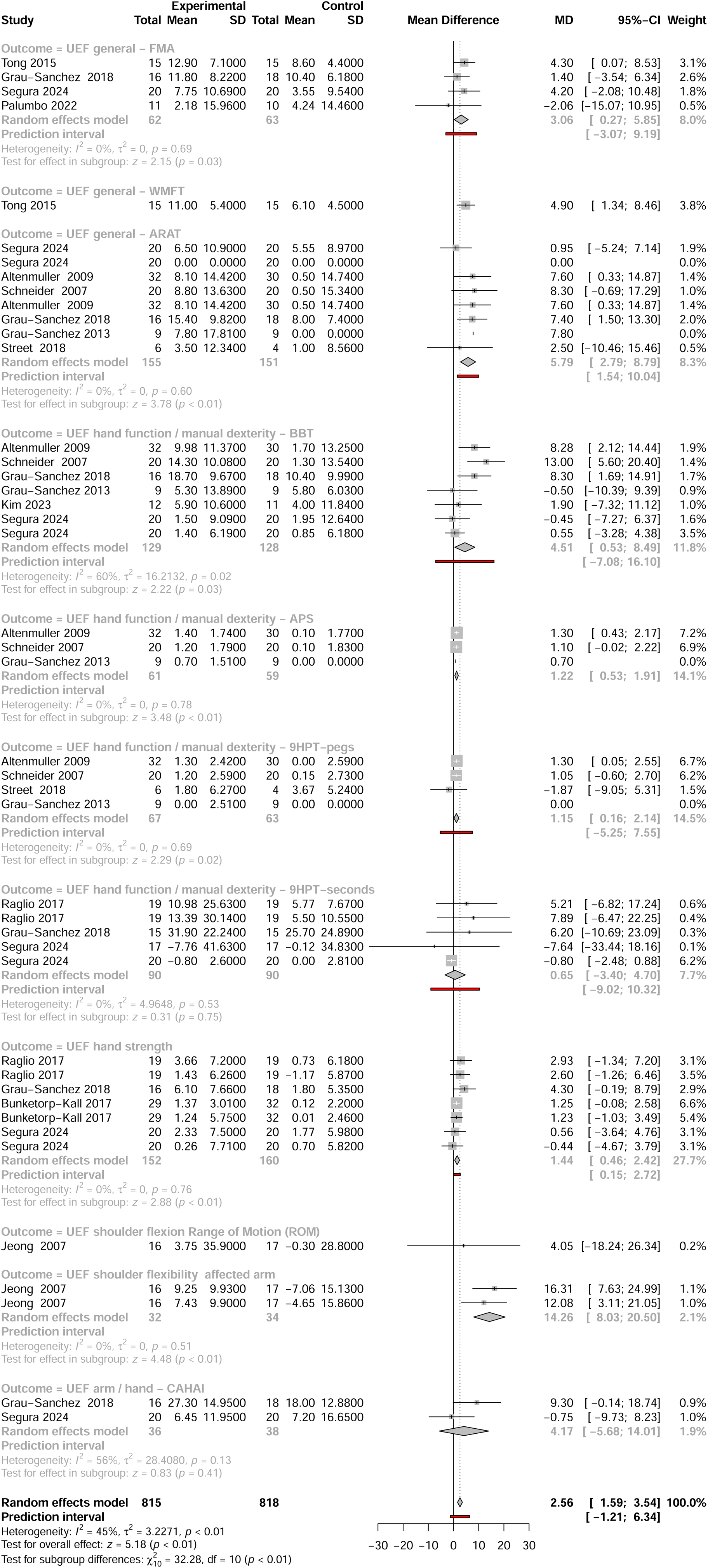
Forest plot of mean differences between experimental and control groups across studies evaluating the effects of intervention on UEF-related outcomes. See legend of Figure 2 for more details.

#### 3.2.2. Language and communication improvements

Sixteen studies reported that MI aid language recovery after BD, with eleven included in a meta-analysis, covering domains like spontaneous speech, repetition, and naming. Across studies, MI consistently outperformed control conditions, particularly for patients with aphasia, as demonstrated in well-designed RCTs ^31–34^.

Enhancements were observed in repetition, articulation, prosody, verbal memory, and general communication. Neuroimaging also revealed increased language-related brain connectivity post-MI ^35^, and intensive singing improved speech-motor functions in nonfluent aphasia ^36^.

A meta-analysis (**Figure 4**) showed significant benefits (MD=3.41, 95% CI: 0.98–5.85; z=2.75, *P*<0.01), despite substantial heterogeneity (I^2^=74%). Repetition and naming (both using AAT) were most responsive, with large, significant effects and no heterogeneity (MD=8.85 and 7.68, respectively; *P*<0.01). General communication and spontaneous speech showed variable results, possibly due to differing tools and baseline impairments. Prediction intervals indicated potential for both positive and null outcomes, highlighting the importance of individual factors in treatment planning.

**Figure 4.**
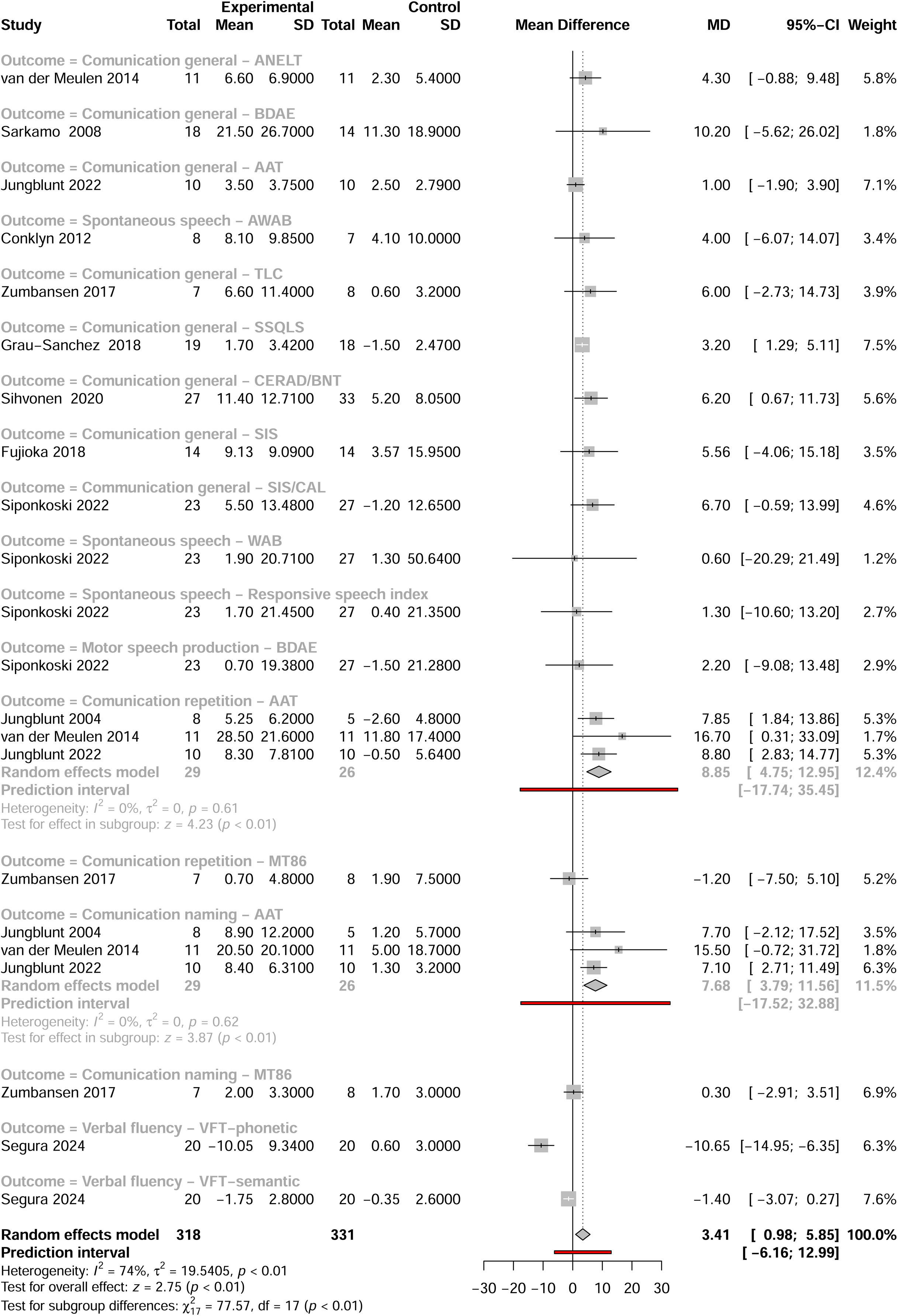
Forest plot of mean differences between experimental and control groups across studies evaluating the effects of intervention on communication-related outcomes. See legend of Figure 2 for more details.

#### 3.2.3. Cognitive rehabilitation

Cognitive deficits are common after BD ^37^. Out of 70 studies reviewed, 21 reported cognitive benefits of MI: 9 on memory, 10 on attention, and 12 on executive function. Music has been shown to aid memory through autobiographical recall ^38^, enhanced brain connectivity ^39^, and verbal memory recovery via vocal music ^40^. Active engagement (e.g., piano playing) supports cortical plasticity, improving attention and executive function in mild TBI ^41^. MI also improves executive function, particularly through instrument playing ^42^, singing ^33^, and improvisation ^43^. Notably, Sihvonen et al. ^44^ showed that Neurological Music Therapy induces white matter plasticity post-TBI. Attention improvements have been noted with musical attention training ^45^, and VR-based music interventions showed promising effects ^46^.

A meta-analysis of 12 studies (**Figure 5**) found a significant overall cognitive benefit (MD=1.11, 95%CI: 0.63−1.59; z=4.52, *P*<0.01), with no observed heterogeneity. Verbal memory (RAVLT) showed a small-to-moderate effect (MD=0.91, *P*<0.01), and visual memory (WAIS-III) had a larger effect (MD=2.58, *P*<0.01). Executive function (TMT-B) showed robust gains (MD=14.42, *P*=0.02). These results confirm the efficacy of MI in cognitive rehabilitation across multiple domains.

**Figure 5.**
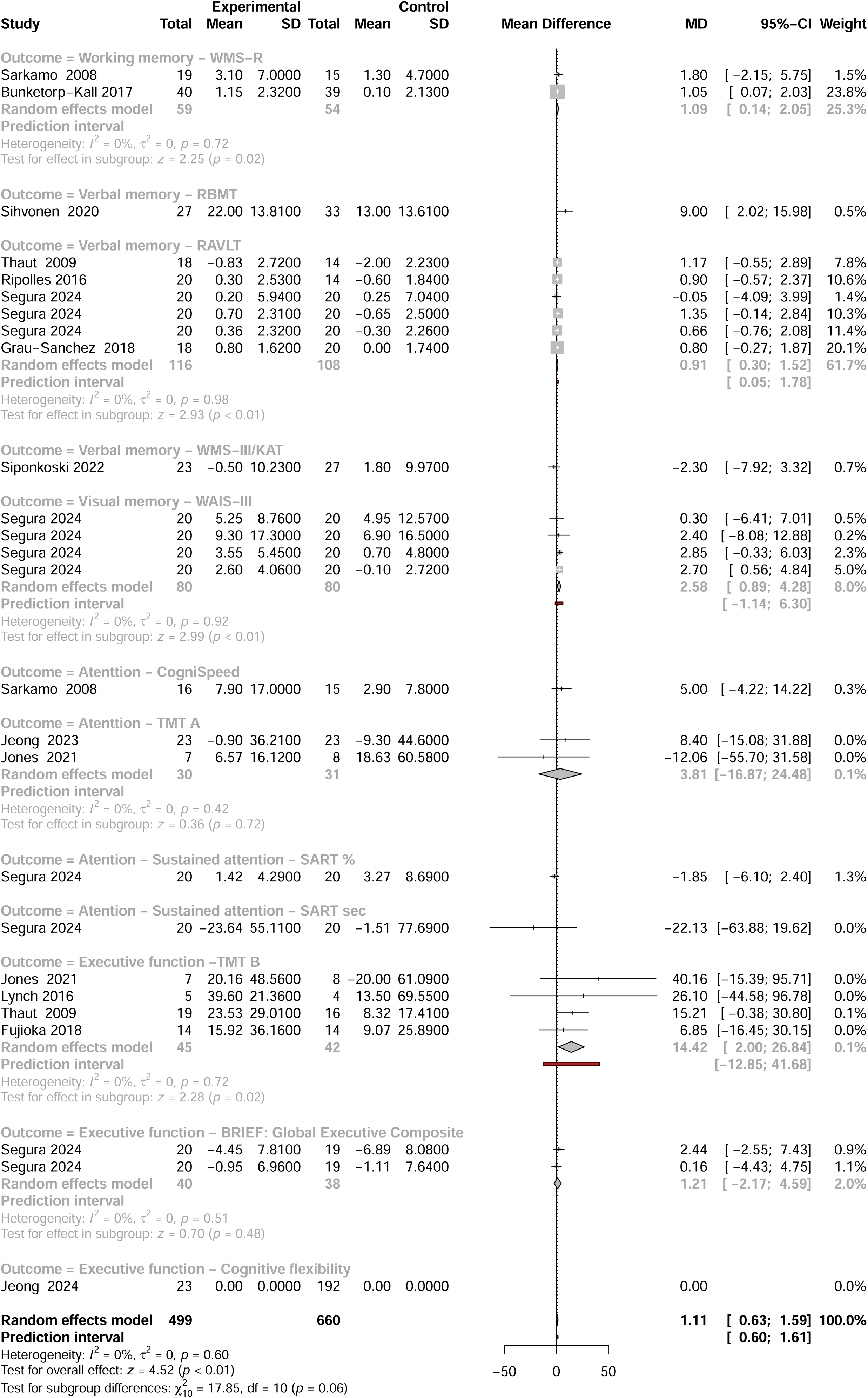
Forest plot of mean differences between experimental and control groups across studies evaluating the effects of intervention on cognitive-related outcomes. See legend of Figure 2 for more details.

#### 3.2.4. Emotional, behavioral, and social outcomes

Twenty-five studies reported emotional benefits from MI, and 24 noted behavioral or social improvements, reduced agitation, improved mood, and reduced depression ^47,48^. Activities such as songwriting, singing, and improvisation enhanced emotional stability ^31,49^. MI reduced agitation in TBI ^50^ and post-stroke depression ^51^. Neuroimaging showed increased consciousness in vegetative or minimally conscious patients exposed to music ^52^. Music also promoted sleep and relaxation ^53,54^.

Biofeedback confirmed music’s relaxing effects ^55^, while fMRI studies linked MI with activation of emotional processing areas ^40^. Social benefits, such as improved interaction, were reported in fewer studies ^41,56^.

A meta-analysis across 12 emotional outcomes (**Figure 6**) showed a small but significant benefit (MD=0.20; 95% CI: 0.04−0.36; z=2.46; *P*=0.01), with moderate heterogeneity (I^2^=49%). Subgroup differences were not significant. Depression-related outcomes were mixed; PHQ-9 and BDI-II showed positive effects; CES-D did not. Emotional well-being improved in some studies (e.g., ^51^), though other measures had wide confidence intervals. Despite some inconsistency, the findings support MI’s emotional and social benefits in BD.

**Figure 6.**
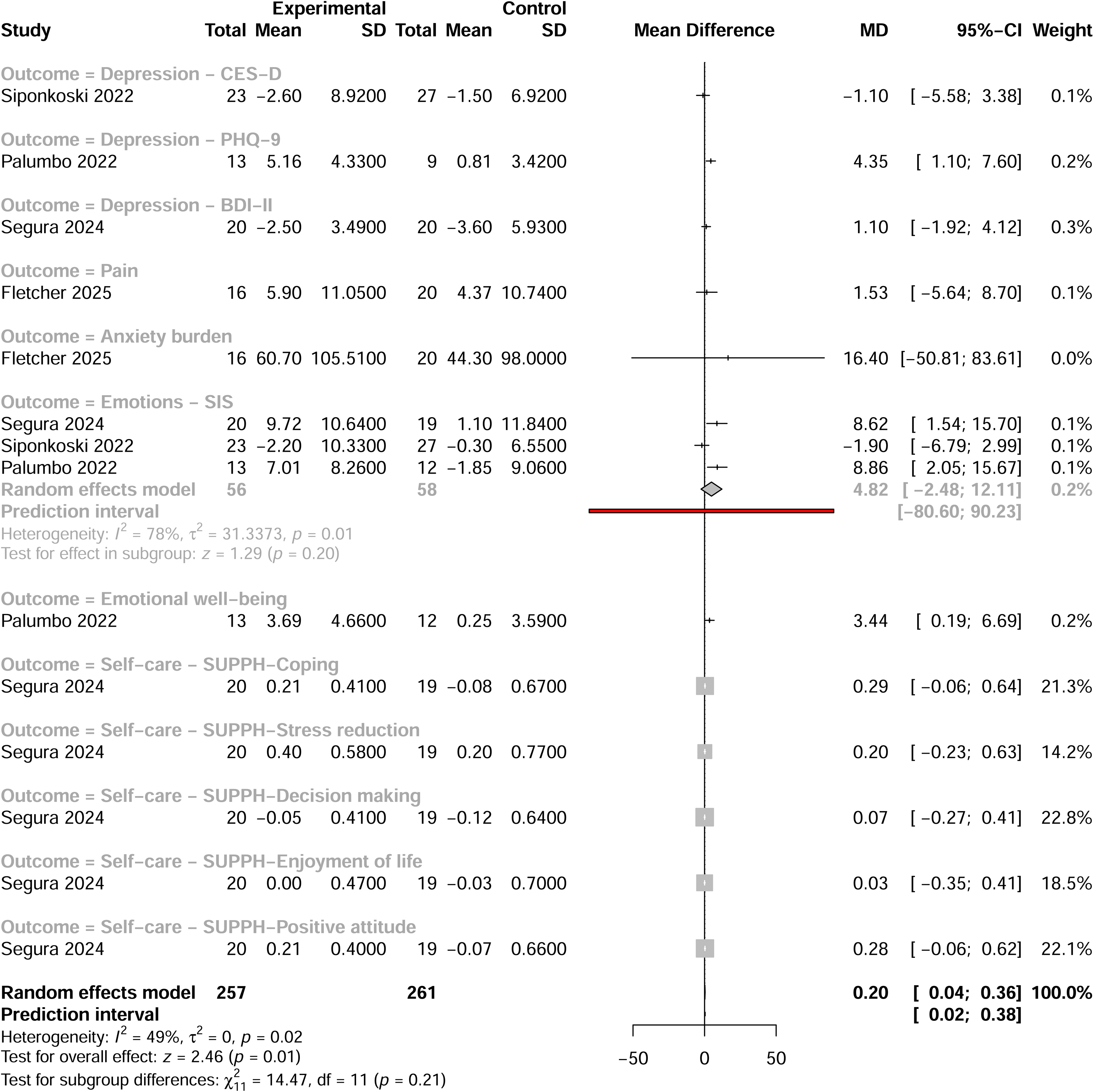
Forest plot of mean differences between experimental and control groups across studies evaluating the effects of intervention on emotional, behavioral and social outcomes. See legend of Figure 2 for more details.

#### 3.2.5. Quality of life

Some studies linked MI to improved QoL, including psychological, physical, and social aspects ^45,48^. A meta-analysis of five studies (**Figure 7**) using three QoL measures (SS-QoL, QoLIBRI, and QoLI) showed a non-significant trend favoring MI (MD=4.44; 95% CI: -1.56–10.43; z=1.45; *P*=0.15), with high heterogeneity (I^2^=73%). Among SS-QoL studies, Cha et al.^25^ reported a strong effect, while others showed modest or no gains. The overall trend suggests possible QoL improvements, but further research is needed to confirm domain-specific effects.

**Figure 7.**
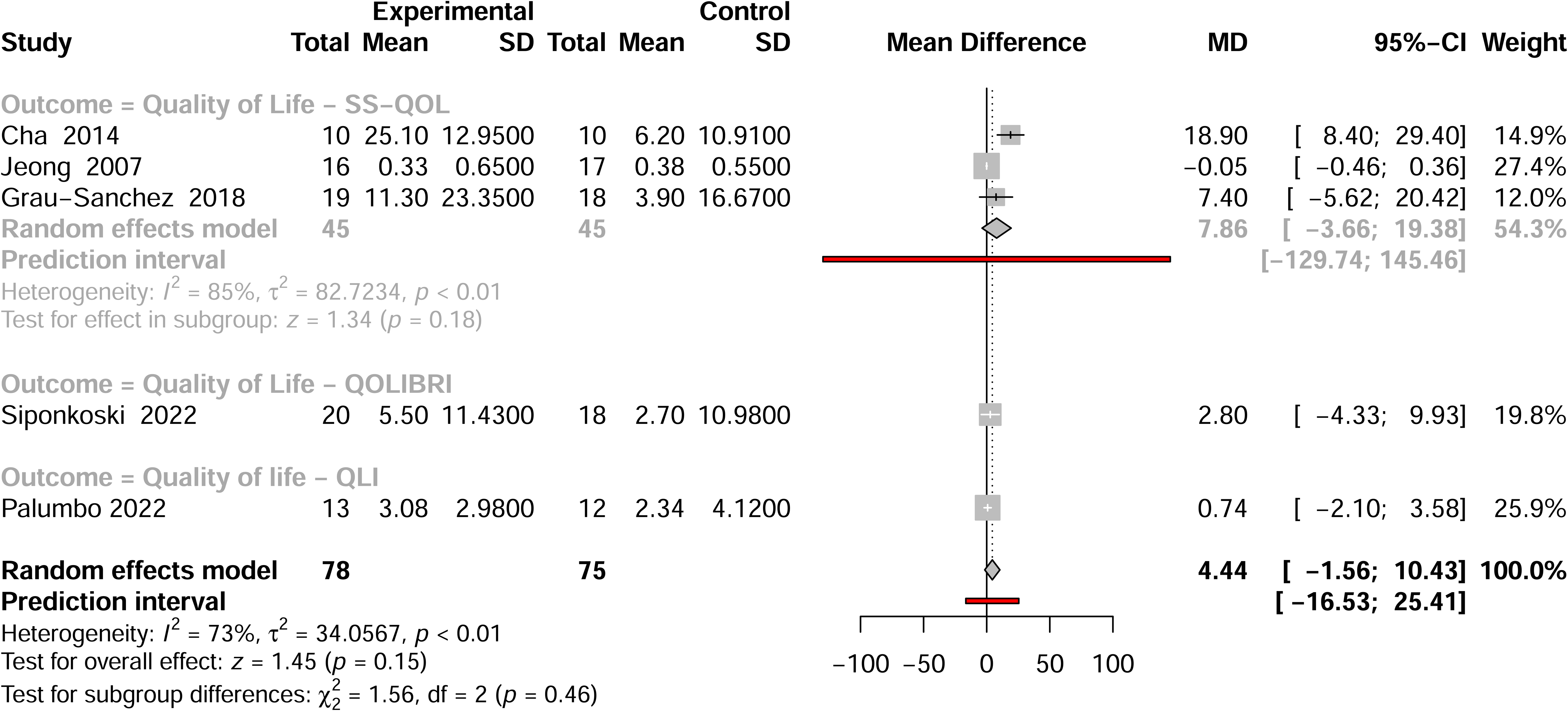
Forest plot of mean differences between experimental and control groups across studies evaluating the effects of intervention on QoL-related outcomes. See legend of Figure 2 for more details.

#### 3.2.6. Global improvement

Global improvement has recently emerged as a relevant outcome in music-based rehabilitation for ABI ^46,51,57^, though earlier reviews did not assess it ^58^. Definitions vary, with studies using cognitive (CDR, GDS), functional (FIM, mRS), or self-reported (SIS) measures.

Meta-analysis of four studies (**Figure 8**) found no significant overall effect of MI (MD=0.03; 95%CI: -0.34–0.41; z=0.18; *P*=0.86; I^2^=31%). Subgroup differences were also non-significant (*χ*^2^=6.61, *P*=0.16), though the SIS Recovery domain showed a marginally positive trend (MD=5.19; 95%CI: -0.42–1.80; z=1.81; *P*=0.07; I^2^=0%). While current evidence is inconclusive, SIS Recovery warrants further study.

**Figure 8.**
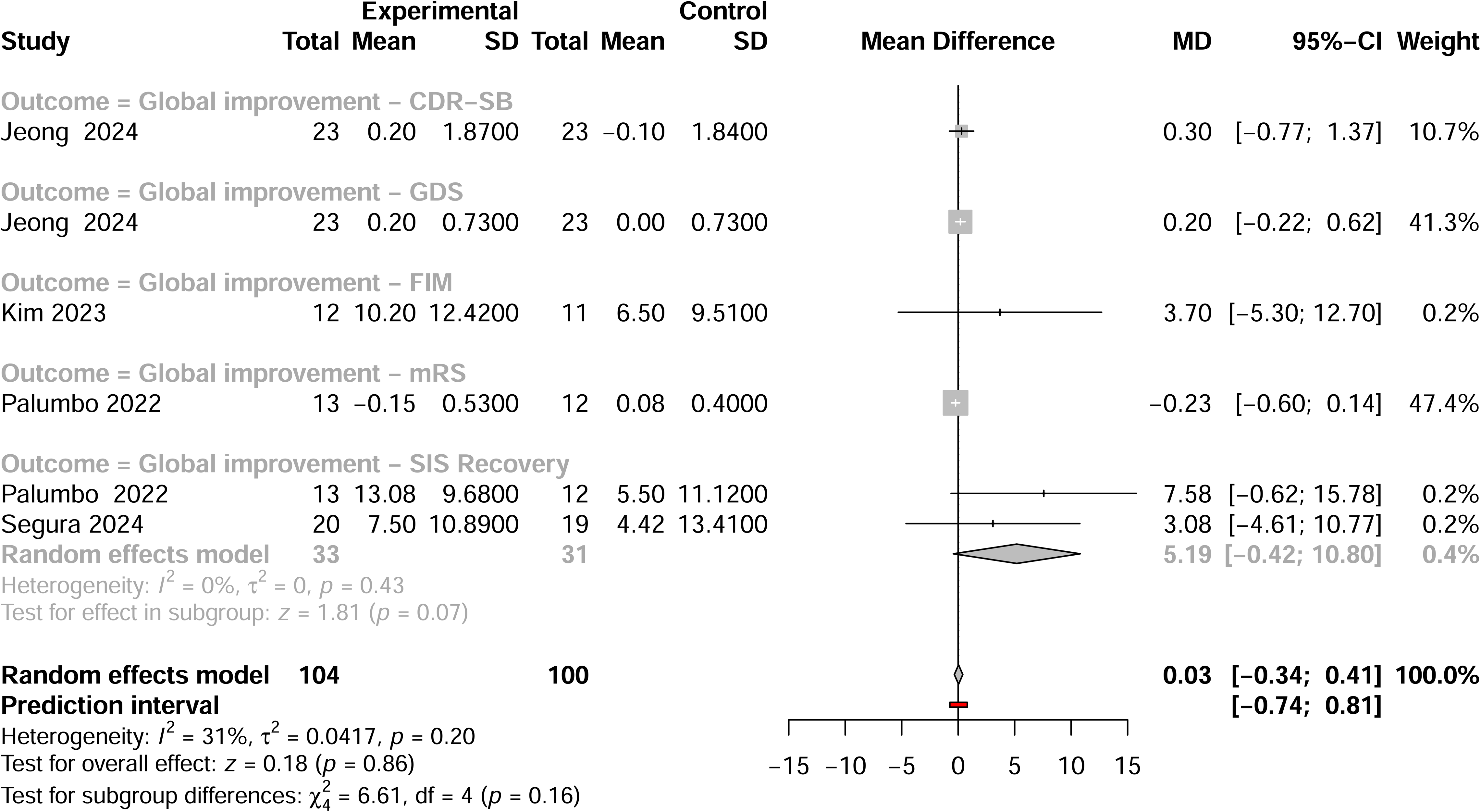
Forest plot of mean differences between experimental and control groups across studies evaluating the effects of intervention on global improvement outcomes. See legend of Figure 2 for more details.

### 3.3. Types of musical interventions in brain damage studies

This review analyzed 70 studies exploring MI for ABI, focusing on their types and associated outcomes. Most interventions were short-term, but seven long-term programs (≥30 hours) consistently showed benefits across cognitive, motor, and emotional domains. These included MST ^30,48^, rhythm- and music-based therapy ^59^, choral singing ^49^, and structured music listening ^31,40^. Active participation, instrument playing, or singing, was most frequent in effective studies (**Table 1**).

**Table 1.** Types of musical interventions (MI) in studies on brain damage (BD). Abbreviations for MI are as follows: FTAS: Fast-tempo auditory stimulation; MACT: Music Attention Control Training; MIT: Melodic Intonation Therapy; MULT-I: Music Upper Limb Therapy-Integrated; NMT: Neurological Music Therapy; MST: Music-supported Therapy; RAMT: Relational Active Music Therapy; RAS: Rhythmic Auditory Stimulation; RMT: Rhythm- and Music-Based Therapy; SIPARI: Singing, Intonation, Prosody, Breathing, Rhythm, Improvisation; VR-MAT: Virtual Reality-based Music Attention; TE: Time Exposure in Hours. Abbreviations for benefits: A: Attention; BS: Behavioral and Social; C: Communication; CF: Cognitive Functions; EF: Executive Function; G: Gait; ME: Memory; EW: Emotional wellbeing; U: UEF; P: PAIN. (*: unclear, one session is reported; **: not specified; ***: with guitar accompaniment; ****: Radio/instrumental classical music/instrumental relaxing music with nature sounds).

| Reference | Musical intervention | TE | Benefits |
| --- | --- | --- | --- |
| ***Interventions involving musical instrument playing*** |  |  |  |
| Chong et al. (2014) | Play the piano or keyboard | 5 | U |
| Bernardi et al. (2017) | Play the piano or keyboard | 2 | BS/CF/U |
| Vik et al. (2018) | Play the piano or keyboard | 4 | A/BS/CF/EF/ME/EW |
| Villeneuve and Lamontagne (2013) | Play the piano or keyboard | 9 | U |
| Villeneuve et al. (2014) | Play the piano or keyboard | 9 | U |
| Kim et al. (2023) | Play the piano or keyboard | 4 | U |
| Altenmuller et al. (2009) | MST | 7.5 | U |
| Schneider et al. (2007) | MST | 7.5 | U |
| Schneider et al. (2010) | MST | 7.5 | U |
| Amengual et al. (2013) | MST | 10 | U |
| Grau-Sánchez (2013) | MST | 10 | BS/U |
| Tong et al. (2015) | MST | 10 | U |
| Ripollés et al. (2016) | MST | 10 | A/BS/CF/EF/EW/U |
| Grau-Sánchez et al. (2018) | MST | 10 | BS/C/EW |
| Rojo et al. (2011) | MST | 20 | G/U |
| van Vugt et al. (2014) | MST | 5 | EW/U |
| Fujioka et al. (2018) | MST | 30 | BS/CF/EF/EW/U |
| Grau-Sánchez et al. (2017) | MST | 18 | U |
| Segura et al. (2021) | MST | 30 | U |
| Segura et al. (2024) | e-MST | 40 | U/ CF |
| Martinez-Molina et al. (2021) | NMT | 20 | CF/EF |
| Lynch et al. (2016) | NMT | 6 | CF/EF |
| Sihvonen et al. (2022) | NMT | 24 | CF/EF |
| Street et al. (2018) | NMT | 6 | U |
| Siponkoski et al. (2020) | NMT | 24 | BS/CF/EF |
| Siponkoski et al. (2022) | NMT | 24 | BS/EW |
| Vaudreuil et al. (2025) | NMT | 8 | EW |
| Raghavan et al. (2016) | MULT-I | 9 | EW/U |
| Palumbo et al. (2022) | MULT-I | 9 | EW/U |
| Raglio et al. (2017) | RAMT | 10 | G/EW/U |
| Jeong et al. (2024) | VR-MAT | 20 | A/EF |
| ***Interventions combining music and movement*** |  |  |  |
| Thompson et al. (2021) | RAS | 6 | G |
| Jeong et al. (2007) | RAS | 16 | BS/EW/U |
| Sheridan et al. (2021) | RAS | 4.5 | G |
| Thaut et al. (2007) | RAS | 15 | BS/G |
| Cha et al. (2014) | RAS | 7.5 | BS/G |
| Park et al. (2010) | FTAS | 10 | G |
| Bunketorp-Käll et al. (2017) | RMT | 36 | G/M/U |
| Mazhari-Jensen et al. (2023) | An exercise program while listening music | 1.5 | G |
| Scholtz et al. (2016) | Sonification | 5 | U |
| Nikmaram et al. (2019) | Sonification | 15 | U |
| Peyre et al. (2023) | Sonification | 3 | U |
| ***Interventions based on singing*** |  |  |  |
| Tamplin et al. (2013) | Choir | 40 | BS/C/EW |
| Zumbansen et al. (2017) | Choir | 34 | C/EW |
| Conckling et al. (2012) | Modified-MIT | <1 | C |
| Bitan et al. (2018) | MIT | 24 | C |
| van der Muelen et al. (2014) | Intensive-MIT | 30 | C |
| Siponkoski et al. (2022b) | MIT | 48 | C |
| Ueda et al. (2024) | MIT | 3.3 | C |
| Marchina et al. (2023) | MIT | 112.5 | C |
| Jungblut et al. (2022) | Voice training SIPARI | 24 | BS/C/CF/EF |
| Baker et al. (2005) | Favorite song*** | 12.5 | C/EW |
| Bower et al. (2014) | Favorite song*** | 2.5 | BS |
| ***Interventions involving passive music listening*** |  |  |  |
| Ribero et al. (2014) | Relaxing music**** | 6 | BS |
| Park et al. (2016) | Classical relax music/preferred music | 9 | BS/EW |
| Kasdan and Kiran et al. (2018) | Musical exercise | 1 | CF/C |
| Sarkamo et al. (2008) | Favorite music | 60 | A/C/CF/ME/EW |
| Sarkamo et al. (2014) | Favorite music | 60 | A/C/CF/ME/EW |
| Carriere et al. (2020) | Favorite music | <1 | BS/C/CF/ME/EW |
| Sihvonen et al. (2020) | Popular/jazz/classical and film music | 60 | C/CF/ME/EW |
| Ushasree et al. (2021) | Traditional songs (Indian ragas) | <1 | BS |
| Baird and Samson et al. (2014) | Popular songs | * | CF/ME |
| Okumura et al. (2014) | 3 clips: beat/instrumental/vocal | 0.03 | BS |
| Fletcher et al. (2025) | Popular/jazz/classical/ relaxing | <6 | P, EW |
| Palmquist et al. (2025) | Relaxing music (classical, jazz, new age) | 5 | P, EW |
| ***Other active and combining musical activities*** |  |  |  |
| Jones et al. (2021) | MACT listening and playing | 3 | A/CF |
| Baker et al. (2019) | Therapeutic songwriting | 12 | EW |
| Seibert et al. (2000) | Listening and playing | ** | A/BS/C/CF/EF/ME/EW |
| Nayak et al. (2000) | Mixed music therapy techniques | 10 | BS/EW |
| Thaut et al. (2009) | Singing and playing improvised music | 2 | CF/EF/EW |

#### 3.3.1. Instrument playing

Instrument-based interventions were applied in 31 studies, targeting fine and gross upper-limb motor skills. Of these, 21 reported significant UEF improvements, particularly through piano-based training ^27,57^. Beyond motor recovery, cognitive enhancements were noted, including in executive functions ^41,60^. MST, involving MIDI keyboards and electronic drums, appeared in 14 studies ^28,61,62^, showing robust motor gains. Neurologic Music Therapy (NMT) also supported motor recovery in seven studies ^42,63^.

#### 3.3.2. Music and movement

Seven studies applied rhythmic auditory techniques like RAS and FAST, improving gait and coordination ^24,26^. Five studies reported improved UEF through rhythm-enhanced movement exercises. Musical sonification of arm movements was explored in three studies ^64,65^, with promising, though preliminary, results.

#### 3.3.3. Singing-based interventions

Singing therapies, used in 11 studies, enhanced speech and communication, especially in aphasic patients. These included choir participation ^49^, Melodic Intonation Therapy (MIT) ^34,66^, and structured vocal training. Particularly, MIT proved more effective than traditional speech-language therapies ^67^.

#### 3.3.4. Passive music listening

Twelve studies focused on recorded music exposure, ranging from classical to patient-preferred music ^31^ ^68^. Benefits included emotional well-being ^69^, improved memory ^40^, and enhanced behavior or communication ^53,70^. Vocal music appeared particularly beneficial for cognitive-linguistic recovery post-stroke.

## 4. Discussion

This systematic review and meta-analysis evaluated the therapeutic effects of MI on individuals with BD, revealing consistent benefits across motor, cognitive, communicative, and emotional domains. Motor rehabilitation emerged as a primary area of benefit. Improvements in UEF and gait were among the most frequently reported outcomes. These effects are likely mediated by mechanisms such as rhythmic entrainment, auditory-motor coupling, and reward-based motivation ^13^, which support motor coordination and timing. For the first time in a systematic review on MI for ABI, we directly linked improvements to specific types of musical activities. Interventions involving music and movement or instrument playing were consistently associated with mobility improvements, while singing-based approaches, such as MIT, demonstrated stronger effects on communicative outcomes. This synthesis of MI variability provides valuable insights for tailoring interventions based on the target domain. Language and communication improvements were also observed, particularly in individuals with aphasia. Interventions such as singing and MIT led to enhanced language production and comprehension ^36,67^. These benefits may result from neural reorganization and strengthened perilesional connectivity ^33,39,40^. However, limitations such as short durations, methodological heterogeneity, and small samples restrict the generalizability of these findings. Cognitive improvements were especially strong, with gains in memory, attention, and executive function. Interventions including active music-making, rhythmic movement, and passive listening were linked to behavioral improvements and neuroplastic changes, particularly in frontal and temporal brain regions ^39,40,42^. These findings suggest music can act as a multisensory stimulus promoting higher-order cognitive recovery. Emotional and psychosocial outcomes were more variable. Some studies reported improvements in mood, anxiety, and depression, although effects on QoL were inconsistent. While positive trends were found, few reached statistical significance, possibly due to subjective assessments and small samples. Nonetheless, studies using self-care subscales (e.g., SUPPH) observed promising outcomes related to coping and stress reduction ^51,71^. Social benefits were often indirectly measured but suggested *via* emotional expression improvements, e.g., on SIS-Emotions subscales.

Interestingly, the review also highlighted how BD affects musical abilities. While acquired amusia is common post-injury ^72^, certain musical functions, especially in individuals with prior musical training, appear preserved. This preservation may reflect the neuroprotective effects of lifelong music engagement through reinforcement of auditory-motor-emotional networks ^73^, revealing a complex pattern of both vulnerability and resilience.

Ultimately, several studies have highlighted the potential of transcriptomic analysis to investigate the biological impact of music across various disease contexts ^10,74,75^. This growing multidisciplinary interest underscores music’s potential not only as a therapeutic tool but also as a modulator of gene expression and neuroplasticity. Understanding these mechanisms will be essential to developing precise, evidence-based MI strategies. Overall, the findings support MI as a promising adjunct in neurorehabilitation. Music consistently demonstrates benefits for motor and cognitive recovery, as well as potential for improving communication and emotional well-being. However, significant challenges remain. Variability in patient populations, intervention protocols, and outcome measures limits comparability across studies. Additionally, small sample sizes and the lack of standardized assessment tools hinder the ability to draw robust conclusions through meta-analyses.

To advance the field, future studies should prioritize large-scale, longitudinal trials with standardized neuropsychological and biological outcome measures, including transcriptomic and epigenomic tools, to better understand the mechanisms driving MI efficacy. With more rigorous designs, MI could become a validated, evidence-based component of comprehensive rehabilitation programs for individuals living with the consequences of BD.

## Data Availability

All data produced in the present work are contained in the manuscript

## 5. Acknowledgements

The authors would like to express their appreciation to the study investigators of the Sensogenomics network (sensogenomics.com; Sensogenomics Working Group [Annex]), as well as the nursery and laboratory service at the Hospital Clínico Universitario de Santiago de Compostela, for their invaluable dedication and support. This work was supported by: *i*) GAIN IN607B 2020/08 and IN607A 2023/02, and EUTERPE_adn (Programa de Cooperación Interreg-VI POCTEP; Ref. 0313_EUTERPE_ADN_1_E) (to A.S.), and IIN607A2021/05 (to F.M.-T.), and *ii)* Consorcio Centro de Investigación Biomédica en Red de Enfermedades Respiratorias (CB21/06/00103; to A.S. and F.M.-T.). The funders were not involved in the study design, collection, analysis, interpretation of data, the writing of this article, or the decision to submit it for publication.

## Supplementary Material

**Table S1**. Summary of studies examining the neuroanatomical regions affected by brain damage (BD) and their impact on musical abilities, highlighting both impairments and notable cases of preserved or enhanced function. This table also includes studies investigating the therapeutic effects of musical stimulation in individuals with brain injuries, consistently reporting positive outcomes across cognitive, emotional, and functional domains.

## Sensogenomics Working Group

Antonio Salas Ellacuriaga – PI; Federico Martinón-Torres – PI; Laura Navarro Ramón – Coordinator

### GenPoB/GenVip - Instituto de Investigación Sanitaria (IDIS) (alphabetic order)

Alba Camino Mera, Albert Padín Villar, Alberto Gómez Carballa, Alejandro Pérez López, Alicia Carballal Fernández, Ana Cotovad Bellas, Ana Isabel Dacosta Urbieta, Narmeen Mallah, Ana María Pastoriza Mourelle, Ana María Senín Ferreiro, Andrés Muy Pérez, Antía Rivas Oural, Antonio Justicia Grande, Antonio Piñeiro García, Anxela Cristina Delgado García, Belén Mosquera Pérez, Blanca Díaz Esteban, Carlos Durán Suárez, Carmen Curros Novo, Carmen Gómez Vieites, Carmen Rodríguez-Tenreiro Sánchez, Celia Varela Pájaro, Claudia Navarro Gonzalo, Cristina Serén Trasorras, Cristina Talavero González, Einés Monteagudo Vilavedra, Estefanía Rey Campos, Esther Montero Campos, Fernando Álvez González, Fernando Caamaño Viñas, Francisco García Iglesias, Gloria Viz Rodríguez, Hugo Alberto Tovar Velasco, Irene Álvarez Rodríguez, Irene García Zuazola, Irene Rivero Calle, Iria Afonso Carrasco, Isabel Ferreirós Vidal, Isabel Lista García, Isabel Rego Lijo, Iván Prieto Gómez, Iván Quintana Cepedal, Jacobo Pardo Seco, Jesús Eirís Puñal, José Gómez Rial, José Manuel Fernández García, José María Martinón Martínez, Julia Cela Mosquera, Julia García Currás, Julián Montoto Louzao, Lara Martínez Martínez, Laura Navarro Ramón, Lidia Piñeiro Rodríguez, Lorenzo Redondo Collazo, Lúa Castelo Martínez, Lucía Company Arciniegas, Luis Crego Rodríguez, Luisa García Vicente, Manuel Vázquez Donsión, María Dolores Martínez García, María Elena Gamborino Caramés, María Elena Sobrino Fernández, María José Currás Tuala, María Martínez Leis, María Soledad Vilas Iglesias, María Sol Rodriguez Calvo, María Teresa Autran García, Marina Casas Pérez, Marta Aldonza Torres, Marta Bouzón Alejandro, Marta Lendoiro Fuentes, Miriam Ben García, Miriam Cebey López, Montserrat López Franco, Nour El Zahraa Mallah, Narmeen Mallah, Natalia García Sánchez, Natalia Vieito Perez, Patricia Regueiro Casuso, Ricardo Suárez Camacho, Rita García Fernández, Rita Varela Estévez, Rosaura Picáns Leis, Ruth Barral Arca, Sandra Carnota Antonio, Sandra Viz Lasheras, Sara Pischedda, Sara Rey Vázquez, Sonia Marcos Alonso, Sonia Serén Fernández, Susana Rey García, Vanesa Álvarez Iglesias, Victoria Redondo Cervantes, Vanesa Álvarez Iglesias, Wiktor Dominik Nowak, Xabier Bello Paderne, Xabier Mazaira López

### Nursing volunteers (alphabetic order)

Alejandra Fernández Méndez, Ana Isabel Abadín Campaña, Ana María León Caamaño, Ana María Buide Illobre, Ángeles Mera Cores, Carmen Nieves Vastro, Carolina Suarez Crego, Concepción Rey Iglesias, Cristina Candal Regueira, Dolores Barreiro Puente, Elvira Rodríguez Rodríguez, Eugenia González Budiño, Eva Rey Álvarez, Fernando Rodríguez Gerpe, Gemma Albela Silva, Isabel Castro Pérez, Isabel Domínguez Ríos, José Ángel Fernández de la Iglesia, José Cruces Vázquez, José Luis Cambeiro Quintela, José Ramón Magariños Iglesias, Julia Rey Brandariz, Julio Abel Fernández López, Luisa García Vicente, Manuel González Lito, Manuel González Lijó, Manuela Pérez Rivas, Margarita Turnes Paredes, María Aurora Méndez López, María Begoña Tomé Arufe, María Campos Torres, María del Carmen Baloira Nogueira, María del Carmen García juan, María Esther Moricosa García, María Luz Chao Jarel, María Martínez Leis, María Mercedes Jiménez Santos, María Salomé Buide Illobre, María Victoria López Pereira, Mercedes Jorge González, Mercedes Isolina Rodríguez Rodríguez, Miren Payo Puente, Natalia Carter Domínguez, Olga María Reyes González, Pilar Mera Rodríguez, Purificación Sebio Brandariz, Salomé Quintáns lago, Yolanda Rodríguez Taboada, María Pereira Grau.

### Other volunteers (alphabetic order)

Alba Arias Gómez, Alejandro Moreno Díaz, Ana Arca Marán, Astro González Guirado, Brais García Iglesias, Carlos Sánchez Rubín, Carmen Otero de Andrés, Clara Pérez Errazquin Barrera, Claudia Rey Posse, Cristina Rojas García, Eduardo Xavier Giménez Bargiela, Elena Gloria Morales García, Fabio Izquierdo García Escribano, Gabriel Guisande García, Jaime López Martín, Lara Pais Ramiro, Lucía Rico Montero, Luís Estévez Martínez, Manuel Estévez Casal, María Aránzazu Palomino Caño, María Rubio Valdés, Marisol Nogales Benítez, Miryam Tilve Pérez, Nuria Villar Muiños, Pablo Del Cerro Rodríguez, Pablo Pozuelo Martínez Cardeñoso, Salma Ouahabi El Ouahabi, Santiago Vázquez Calvache

