## Supplementary Text 1 for "Systematic review and meta-analysis reveal positive therapeutic effects of music in brain damage rehabilitation"

**Detailed Method’s section**

Methodological aspects of the systematic review

A systematic review was conducted using the PubMed database, with the initial search performed up to March 24, 2023. To ensure comprehensive coverage of literature relevant to BD, we selected search terms such as "brain injur*" (which considers both terms, injury and injuries), "brain deformation", and "cerebral damage", based on their frequency and relevance in the literature. The term "music" was intentionally kept broad to avoid limiting the scope of the study. We applied the [tiab] field tag (title and abstract) to restrict results to articles containing these specific terms, thereby filtering out irrelevant publications. The final search query was as follows: ("brain injur*"[tiab] OR "brain damage*"[tiab] OR "brain deformation*"[tiab] OR "cerebral injur*"[tiab] OR "cerebral damage*"[tiab]) AND (music*[tiab]).

This initial search yielded 247 records. These were refined by removing duplicates, systematic reviews, and meta-analyses using PubMed’s built-in filters. Additional filters were applied to include only English-language studies focused on human subjects, published between 2000 and the present. This resulted in a total of 92 publications. Of these, 13 were excluded for being reviews or systematic reviews. The remaining 79 articles underwent detailed screening, during which we excluded studies that were not directly related to the impact of music on Acquired Brain Injury (ABI), as well as those combining music therapy with other simultaneous interventions. We also excluded one article focused on congenital brain disorders, as this review specifically targets ABI.

Following this process, 23 publications met all inclusion criteria and were selected for the systematic review. Additionally, 34 relevant articles were incorporated through reference mining of previous reviews and other key sources.

To ensure the relevance and completeness of the review, a second search was conducted on June 11, 2025. This search extended to multiple databases, including PubMed, Embase, Scopus, the Cochrane Library Core Collection, Web of Science, and ClinicalTrials.gov. The same search terms and strategy were used to maintain consistency, with filters applied to restrict results to studies published between 2023 and 2025. This yielded a total of 311 results across all platforms. Using the automation tool Rayyan (ryyan.ai), 157 duplicates were automatically removed, and one additional duplicate was removed manually, resulting in 153 unique articles. These were screened manually, leading to the exclusion of 145 records primarily for not focusing on BD, MI, or lacking an experimental design (**Figure 1**). Of the remaining articles, nine were retrieved for full-text review, with one article ultimately not accessible. Following expert consultation, four recently published studies were added, bringing the final total to 70 publications included in the systematic review.

This review was conducted and reported in agreement with the Preferred Reporting Items for Systematic Reviews and Meta-Analyses (PRISMA) guidelines (https://www.bmj.com/content/372/bmj.n71); see **Figure 1**.

Meta-analysis

Complementing the qualitative synthesis, this review incorporates a meta-analysis of 31 studies that provided complete data suitable for statistical evaluation. The analysis targets key outcomes frequently assessed in BD research, including general improvement, motor recovery, communication, cognitive rehabilitation, QoL, and emotional well-being. A standardized approach was used to synthesize findings across these diverse domains to ensure methodological rigor. Data extraction was conducted independently by two reviewers. Studies were excluded if they lacked a control group, focused exclusively on acute physiological outcomes, or did not isolate the effects of music from other concurrent interventions.

Extracted data included sample sizes, means, and standard deviations for both experimental and control groups, type of outcome measure used, intervention duration, and study design characteristics. For each outcome, we estimated the pooled mean difference and corresponding 95% confidence intervals (CIs) in change scores between baseline and follow-up, comparing the intervention group to non-interventional controls. This was performed using a random-effects meta-analysis, based on the DerSimonian and Laird method ^1^. Although a random-effects model yields wider CIs (accounting for uncertainty due to between-study variability), it is the recommended approach when heterogeneity is moderate to high (as confirmed in several analyses in this study, with I^2^ > 50% in some domains) or when studies differ significantly in design, participant characteristics, and intervention methods. Given the considerable heterogeneity in musical intervention procedures for brain-damaged populations, the random-effects model provides a more appropriate and reliable estimation of overall effects. Therefore, its use enhances the validity and generalizability of the findings across diverse clinical contexts.

Between-study heterogeneity was evaluated using the I^2^ statistic, tau-squared (τ^2^), and Cochran’s Q test. Where substantial heterogeneity was present (I^2^ > 50%), subgroup analyses were conducted to explore potential sources, and subgroup differences were tested using chi-squared (χ^2^) statistics. Forest plots were generated to display both individual study outcomes and pooled effect estimates. Although the limited number of included studies precluded formal tests for publication bias (e.g., funnel plots or Egger’s regression), sensitivity analyses were conducted. These included removing studies with outlier results or high risk of bias to assess the robustness of the pooled findings. In addition to overall estimates, subgroup analyses were also stratified by outcome type (e.g., Fugl-Meyer Assessment [FMA], Action Research Arm Test [ARAT], Box and Block Test [BBT]), reflecting the multidimensional nature of e.g., Upper Extremity Function (UEF) assessments and identifying scale-specific variations in intervention effects.

Prediction intervals were also computed to estimate the dispersion of musical treatment impacts in new study settings, accounting for both within- and between-study variance.

A common challenge in BD research is the frequent absence of reported standard deviations for change-from-baseline scores ^2^. Following Cochrane guidelines ^3^, we imputed missing standard deviations using a conservative correlation coefficient of *r* = 0.8, consistent with assumptions adopted in previous comparable meta-analyses ^2,4^.

To ensure consistency in the direction of treatment effects, outcome scores were harmonized by reversing the direction of specific scales when necessary. When studies used different instruments to assess the same outcome (e.g., UEF), scores were standardized or converted to a common metric to facilitate meaningful comparison. However, due to differences in scale types, scoring conventions, and units, some studies could not be included in joint subgroup analyses, limiting the comparability within certain subdomains. Despite this limitation, the global pattern of results remained robust and informative.

Accordingly, separate meta-analyses were performed for each functional domain (e.g., UEF, gait, cognition), ensuring all effect sizes were aligned and interpretable. All statistical analyses were conducted using the META package in R ^5^, and forest plots were used to visualize the distribution of effect sizes. The statistical significance of pooled effects was assessed using the z-test.
