## Supplementary Text 2 for "Systematic review and meta-analysis reveal positive therapeutic effects of music in brain damage rehabilitation"

**Extended results section – Meta-analysis**

**1. Bibliographic systematic review and meta-analysis**

Acquired Brain Injury (ABI) refers to the brain damage that occurs after birth due to external factors, such as Traumatic Brain Injury (TBI), or internal factors like stroke, aneurysm, tumor, infectious disease, or heart attack (non-TBI) ^1^. The diversity of brain injuries poses a significant challenge for music-based studies aiming to draw conclusive results regarding the scope and connections between music and these biological conditions. For instance, among the reviewed studies, we identified 15 articles focusing on TBI patients, 39 on non-TBI patients, and some targeting specific diagnoses like aphasia, alexia, agnosia, amusia, or Diffuse Brain Injury (DBI), Right Brain Damage (RBD), and Left-Brain Damage (LBD), or in ABI in general. The number of participants varied considerably over the studies, with the majority involving small sample sizes and only a few including more than 50 participants. This diversity can be attributed to the inherent heterogeneity of the ABI population and the challenges of enrolling and managing large groups of patients with BD in musical activities. All identified studies fall within the fields of neuroscience and psychophysiology, comprising experimental research that investigates the restorative effects of music as a form of rehabilitation for individuals with BD (see **Section 3.2**). To further this analysis, we conducted a meta-analysis focusing on the types of music-based interventions used in these experimental studies. This approach allowed for a deeper exploration of the diverse applications of music as both a clinical tool and a subject of research in therapeutic contexts (see **Section 3.3**).

**2. The therapeutic potential of music training in neurorehabilitation**

The therapeutic potential of music training is well supported by scientific evidence, yet its clinical implementation remains limited. Since 2000, a total of 70 studies have examined the effects of musical stimulation in individuals with BD, consistently reporting beneficial outcomes across a range of functions (**Table S1**). However, considerable variability exists in study designs, participant profiles, and intervention protocols, which complicates standardization and broader clinical adoption.

The experimental designs of the included studies were heterogeneous. They comprised randomized controlled trials (RCTs), detailed case reports, studies employing neuroimaging to assess functional or structural brain changes, clinical trials, and quasi-experimental designs. Many of these investigations evaluated pre- and post-intervention effects or compared outcomes with alternative therapies or healthy control groups.

Analysis of this body of research reveals that music-based rehabilitation contributes to improvements in behavioral, cognitive, and motor functions, while also promoting neuroanatomical reorganization during recovery. These therapeutic effects are categorized by outcome domains such as motor function, communication skills, cognitive performance, emotional regulation, behavior, and social engagement. To strengthen the evidence base, we conducted a meta-analysis when sufficient data were available in these studies (*n* = 31), targeting specific outcome domains to provide a rigorous and quantitative synthesis of the findings. This approach enables a clearer, more systematic evaluation of music’s rehabilitative efficacy and highlights its potential as a complementary tool in neurorehabilitation.

*2.1. Benefits of motor rehabilitation*

Thirty-two studies have demonstrated the effectiveness of MI in motor rehabilitation for individuals with BD. Since 2000, eight studies have reported improvements in gait parameters, while 26 studies have focused on UEF recovery. In particular, Music-Supported Therapy (MST), a type of MI based on piano and percussion exercises, has gained recognition as a powerful tool for motor rehabilitation, with enhanced mobility emerging as the most consistently reported benefit across the reviewed studies (see also **Table S1**).

2.1.1. Gait functional recovery

Gait rehabilitation is crucial, as psychomotor impairments affecting walking patterns are a common consequence of cerebral damage. The results demonstrated significant improvements across all categories in participants who engaged in MI compared to control groups. Among the most widely used approaches, Rhythmic Auditory Stimulation (RAS) is particularly effective in gait training. RAS involves synchronizing movements with musical rhythms, often enhanced by a metronome to provide structured auditory cues ^2-6^. Studies that excluded musical stimuli from the RAS protocol were not included in this review. Notably, four comparative RCTs demonstrated that rhythmic stimulation therapies are more effective for gait rehabilitation than standard physiotherapy without music ^4-7^.

A meta-analysis of five studies assessed gait-related outcomes, including general gait performance, step length, symmetry, cadence, balance, timing, and velocity (**Figure 2**). This meta-analysis offers robust evidence for the positive influence of MI on various aspects of gait performance in individuals with BD. The pooled analysis revealed a pooled mean difference of 5.47 (95%CI: 2.08–8.86) and a statistically significant overall effect (z-score = 3.16, *P*-value < 0.01), supporting the notion that musical stimulation can facilitate improvements in locomotor function. Nevertheless, the analysis revealed high heterogeneity across studies, with an I^2^ of 96% (τ^2^ = 35.84; *P*-value < 0.01), indicating marked variability in effect sizes. This level of heterogeneity is likely due to a range of methodological differences, including patient populations, intervention durations, outcome definitions, and measured outcomes (different assessment tools). Further reflecting this variability, the subgroup analysis found significant differences across the various gait outcomes assessed (χ^2^ = 135.94, df = 6, *P*-value < 0.01). When examining individual gait domains, several emerged as particularly responsive with strong and statistically significant improvements. Gait length, cadence, and velocity demonstrated clear benefits from MI, with effect sizes that were both positive and meaningful. These outcomes showed particularly strong effects, as indicated by their respective subgroup z-values (z-score = 6.31, z-score = 8.76, and z-score = 7.28, respectively, all with *P*-value < 0.01), and relatively low heterogeneity (I^2^ values: 49% [τ^2^ < 0.0001], 9% [τ^2^ = 2.32], and 26% [τ^2^ = 3.09]; respectively). These results suggest that MI may effectively support temporal and pacing aspects of walking. Conversely, outcomes related to gait balance and symmetry displayed mixed or nonsignificant results, with subgroup z-values of 1.48 (*P*-value = 0.14) for balance general and 1.30 (*P*-value = 0.20) for balance timing, and CIs for the mean differences overlapping zero. This may indicate that while MI readily support timing and stride regularity, more complex postural or symmetrical adjustments may require different or more targeted therapeutic strategies. Importantly, the overall prediction interval remained positive, suggesting that future studies are likely to observe beneficial effects even when accounting for between-study variability. The relatively low heterogeneity observed in some subgroups, such as cadence and velocity, also strengthens the case for these specific metrics as reliable and responsive indicators of progress during music-based gait rehabilitation.

2.1.2. Upper-extremity function (UEF) recovery

Twenty-six studies investigated the impact of musical interventions on UEF recovery, consistently reporting notable benefits. Research in this field suggests MI enhances motor function by engaging individuals in active instrumental training, such as playing the piano keyboard or electric drums, which provides direct feedback and sensory information. According to different authors, MI can improve: *i*) Hand rehabilitation ^8-11^, increasing finger movement velocity and pressing force ^12,13^, *ii*) Shoulder flexibility, and ankle flexion and extension ^14,15^, *iii*) Fine and gross motor skills of the hand and arm, including speed, precision, and smoothness. ^16-19^, *iv*) Functional grasping and pinching movements ^20^, *v*) Neuroplasticity and motor recovery, restoring connectivity between auditory and motor regions ^16,21-23^. In addition, Schneider et al. ^24^ and Segura et al. ^25^ demonstrated that MST is more efficient than conventional physiotherapy for the recovery of fine motor skills in stroke patients.

A comprehensive meta-analysis of 12 studies assessed the impact of MI on UEF across several outcome domains, including general motor performance, manual dexterity, hand strength, and shoulder mobility (**Figure 3**). The meta-analysis consolidates evidence from a broad range of studies assessing the effects of MI on UEF in patients with BD. The overall pooled analysis revealed a clear and statistically significant benefit, with a mean difference of 2.56 (95%CI: 1.59–3.54) and a highly significant test for overall effect (z-score = 5.18, *P*-value < 0.01). This strong signal indicates that, across diverse study designs and outcome measures, musical stimulation contributes positively to motor recovery in affected individuals. This strong effect was accompanied by a favorable prediction interval (-1.21; 6.34), reinforcing the likelihood that future studies will also detect improvements. Heterogeneity was moderate (I^2^ = 45%, τ^2^ = 3.23), reflecting methodological differences among studies, but subgroup differences were significant (χ^2^ = 32.28, df = 10, *P*-value < 0.01), indicating that the effect size varied substantially across functional domains. This variation emphasizes the importance of considering specific functional domains when evaluating the impact of MI. Detailed subgroup analyses shed light on where the most robust effects occur. Seven out of 10 subgroups showed statistical significance in the test for subgroup effect, and all of them with mean differences that are positive. Some of the most prominent and consistent benefits were observed in UEF general, manual dexterity, and shoulder flexibility, which are critical for functional independence in daily life. The Box and Block Test (BBT) scores demonstrated a significant improvement with a mean difference of 4.51 (95%CI: 0.53–8.49; z-score = 2.22, *P*-value = 0.03) in UEF hand function/manual dexterity. Similarly, the Nine-Hole Pegboard Test (9HPT-pegs) scores showed a significant improvement of 1.15 (95%CI: 0.16–2.14; z-score = 2.29, *P*-value = 0.02). Both measures exhibited variable heterogeneity (BBT: I^2^ = 60%; 9HPT-pegs: I^2^ = 0%). These findings strongly suggest that rhythmic and MI provide robust improvements in fine motor control and coordination. Task-oriented functional assessments also showed compelling effects. The Action Research Arm Test (ARAT) produced one of the largest effect sizes (MD = 5.79, 95%CI: 2.79–8.79) with a strongly significant test (z-score = 3.78, *P*-value < 0.01), indicating substantial improvements in complex, goal-directed upper-limb tasks. Global impairment scales such as the Fugl-Meyer Assessment (FMA) showed a statistically significant improvement (z-score = 2.15, *P*-value = 0.03), reinforcing the idea that broader motor impairment measures are highly sensitive to short-term or specific functional changes induced by MI. Other outcomes in the general UEF, such as hand strength and shoulder flexibility, also demonstrated a strong effect, further supporting the conclusion that MI can markedly enhance task-specific motor recovery. These measures, together with dexterity outcomes, represent the most significant and clinically meaningful improvements observed in this meta-analysis.

These findings underline the value of music-based protocols for promoting fine motor control and functional task performance, which are essential for independence in daily activities. The results also highlight a relative limitation in improving strength and joint flexibility, indicating that these domains may require complementary rehabilitation approaches. Particularly in these UEF outcomes, the observations suggest that future research should prioritize standardized protocols and explore the integration of music-based therapy with interventions targeting strength and proximal mobility to maximize overall recovery.

*2.2. Language and communication improvements*

Sixteen studies have reported that musical interventions stimulate language recovery following BD. Eleven of these studies were included in a meta-analysis, covering various language-related domains such as general communication, spontaneous speech, repetition, and naming. The findings consistently demonstrated greater improvements in participants receiving MI compared to control groups. Most of the studies focused on patients with aphasia, highlighting the effectiveness of musical treatments in enhancing communication abilities, particularly in several well-designed randomized controlled trials (RCTs) ^26-32^. Reported improvements included reading and repetition abilities ^26^; articulation, prosody in spontaneous speech, naming, repetition, and comprehension ^30^; verbal memory and overall language recovery ^33^; expressive speaking and vocal range ^34^; as well as general communication and vocalization. Interestingly, neuroimaging studies have also shown increased connectivity in brain regions associated with language following musical intervention ^35^. Additionally, intensive singing has been found to enhance speech motor functions in individuals with nonfluent aphasia after BD ^32,36^.

A meta-analysis of these studies allows the evaluation of the effectiveness of rehabilitative interventions on communication outcomes in individuals with BD resulting from stroke or TBI (**Figure 4**). The pooled results indicate a statistically significant overall benefit of experimental interventions compared to control conditions, with a pooled mean difference of 3.41 (95%CI: 0.98–5.85) and a z-score of 2.75 (*P*-value < 0.01) in the test for overall effect. These findings suggest that these therapies contribute to meaningful improvements in language abilities. However, substantial heterogeneity was observed across studies (I^2^ = 74%, τ^2^ = 19.54, *P*-value < 0.01), likely reflecting variability in intervention types, outcome measures, and participant characteristics. Subgroup analyses revealed that certain communication domains responded more consistently to MI. In particular, outcomes related to communication repetition and naming (AAT) demonstrated robust and statistically significant improvements with high mean differences (repetition MD = 8.85 and 95% of 4.75–12.95; naming MD = 7.68 and 95%CI of 3.79–11.56), with no observed heterogeneity (I^2^ = 0%, τ^2^ = 0, *P*-value > 0.60 for both) and strong effect sizes (z-score = 4.23 and z-score = 3.87, respectively; *P*-value < 0.01 for both). These findings highlight the responsiveness of these specific language functions to structured therapeutic input. General communication outcomes (e.g., ANELT, BDAE, AAT, TLC, SIS, CERAD/BNT) and spontaneous speech showed variable results, with several studies reporting wide CIs and non-significant effects but positive mean differences. This variability may be attributed to differences in assessment tools, sample sizes, and baseline impairment severity. The prediction interval for the overall effect (95%CI: -6.16–12.99) suggests that while future studies may often observe positive outcomes, negative or null effects remain possible, reinforcing the importance of considering patient-level factors, such as aphasia type and severity, when evaluating treatment efficacy. Overall, the findings support the clinical relevance of targeted speech and language MI, especially for repetition and naming abilities, and emphasize the importance of integrating evidence-based approaches in the rehabilitation of acquired communication impairments following BD.

*2.3. Cognitive rehabilitation*

Cognitive impairment is one of the most common consequences of BD ^37,38^. In this review, 21 studies (out of 70 included in this study) reported cognitive benefits following MI, with 9 focusing on memory, 10 on attention, and 12 on executive function.

Several studies have highlighted the role of music in memory recovery. One study demonstrated that listening to popular songs evoked autobiographical memories in five individuals with severe ABI, providing the first evidence of this effect in patients with severe BD ^39^. Other research has shown that listening to favorite music enhances brain connectivity and activates memory-related functions ^27,40^. Moreover, different musical elements appear to influence memory recovery in distinct ways. For example, vocal music has been found to enhance verbal memory recovery more effectively than instrumental music or audiobooks ^33^. Additionally, engaging in active musical activities, such as playing the piano, promotes cortical plasticity by stimulating neural connections, thereby improving memory, attention, and executive function in individuals with mild TBI ^41^.

Deficits in executive functions are considered core symptoms of TBI. Numerous studies suggest that MI effectively stimulates executive function, particularly through active musical engagement. Research has demonstrated that executive function is activated and improved through various musical experiences, including: *i*) Playing musical instruments ^23,41-46^, *ii*) Singing therapy in aphasia recovery ^30^, and *iii*) Musical improvisation ^47^. A particularly compelling study by Sihvonen et al. ^48^ demonstrated that Neurological Music Therapy (NMT), which includes rhythmic training, cognitive-motor training, and piano and drum playing, induces structural white matter neuroplasticity in post-TBI patients, providing a biological basis for improved executive function.

Moreover, selective attention is significantly affected in ABI patients ^49^. MST, which involves active training using electronic drums and a piano keyboard, has demonstrated significant improvements in attention, executive functions, information processing speed, and mental flexibility in individuals with chronic stroke ^23,42^. A recent study by Jeong et al. ^50^ further demonstrated that virtual reality-based music attention training is an effective cognitive intervention for restoring attentional processes in the ABI population.

A meta-analysis on memory, attention, and executive function, incorporating 12 studies, confirms the positive impact of music on cognitive recovery in individuals with BD (**Figure 5**). The meta-analysis included data that allows assessing five key cognitive domains: working memory, verbal memory, visual memory, attention, and executive function. When all cognitive outcomes were pooled together, combining both executive function and general cognition, the analysis yielded a significant overall effect size of 1.11 (95%CI: 0.63−1.59) with strong statistical significance (z-score = 4.52, *P*-value < 0.01). Although some variability was observed between cognitive subgroups, the overall heterogeneity was zero (I^2^ = τ^2^ = 0%, *P*-value < 0.60). The findings provide encouraging evidence for the efficacy of MI in enhancing cognitive outcomes in neurorehabilitation. However, the test for subgroup differences was marginally significant (χ^2^ = 17.85, df = 10, *P*-value = 0.06), suggesting that the magnitude of benefit might differ depending on the specific cognitive domain targeted. In the subgroup analysis, we found statistically significant results in four out of 10 subgroups, all with positive mean differences and strong effects. In the general cognition subgroup, specifically verbal memory outcomes assessed using the RAVLT, the mean difference was 0.91 with CIs consistently showing a positive trend (95%CI: 0.30−1.52). The test for overall effect in this domain was statistically significant (z-score = 2.93, *P*-value < 0.01), indicating a small-to-moderate benefit of MI in cognitive performance. Importantly, heterogeneity within this subgroup was negligible (I^2^ = 0%, τ^2^ = 0, *P*-value = 0.98), suggesting highly consistent results across studies. The visual memory subgroup measured using WAIS-III demonstrated a higher effect, with a pooled mean difference of 2.58 (95%CI: 0.89−4.28), with the overall subgroup result being statistically significant (z-score = 2.99, *P*-value < 0.01) and no heterogeneity across studies (I^2^ = τ^2^ = 0%, *P*-value < 0.92). The results are even more robust in the executive function-TMT B subgroup, with a high and significant mean difference (MD = 14.42; 95%CI: 2.00−26.84) and strong effect (z-score = 2.28, *P*-value = 0.02) and no heterogeneity across studies (I^2^ = τ^2^ = 0%, *P*-value < 0.72), indicating a robust benefit of MI in executive function. This meta-analysis on cognitive rehabilitation provides compelling evidence that MI can significantly enhance cognitive recovery in individuals with BD, particularly in the domains of executive function, verbal memory, and visual memory. The overall large effect size and consistent findings across studies (with minimal heterogeneity) highlight the reliability and robustness of these outcomes. While some domain-specific variability exists, the positive and statistically significant effects across key cognitive areas highlight the potential use of music as an effective tool in cognitive neurorehabilitation for this population.

*2.4. Emotional, behavioral, and social outcomes*

Twenty-five studies have reported improvements in emotional well-being following MI, while 24 studies have documented behavioral and social benefits, including reductions in agitation, enhanced consciousness, increased relaxation, and decreased symptoms of depression, among others ^5,14,21,23,42,51,52^. A qualitative assessment of these studies provides deeper insight into music’s impact on behavioral and emotional regulation in patients with BD. Specifically, 25 studies examined the effect of MI on mood and emotional regulation in BD patients, consistently reporting significant benefits. Musical activities such as songwriting, singing, playing an instrument, choral singing, and improvisation were shown to enhance emotional well-being and mood stability in individuals with ABI [ ^14,23,26,27,34,53-56^.

Music’s ability to reduce agitation emerged as a key outcome in studies involving TBI patients ^55,57^, as did its effect in reducing post-stroke depression ^52^. Furthermore, two neuroimaging studies used MRI scans to observe patients in vegetative or minimally conscious states after BD while they were exposed to music, revealing a positive effect on levels of consciousness ^40,58^ . Another study involving six TBI patients employed EEG readings taken before, during, and after listening to a music raga, demonstrating that music can promote relaxation and deep sleep ^59^. A more recent study also demonstrated the efficacy of music listening in improving sleep during post-acute ABI rehabilitation ^60^. Additionally, a recent case report supported the use of music to reduce anxiety and promote relaxation, as confirmed through biofeedback measures in a TBI patient ^61^. Similarly, Ribeiro et al. ^62^ found that music effectively induced relaxation in 13 individuals with severe cerebral damage in a vegetative state, and Fletcher et al. ^63^ observed positive trends and reduced variability in anxiety and pain in acute stroke patients over 24 hours. Several studies have confirmed that musical stimulation activates brain regions involved in emotional processing ^33,40,41^, providing neurobiological evidence of music’s role in emotional regulation and mood enhancement.

The social benefits of MI have been explored in a smaller number of studies, which reported improvements in social interaction and communication skills after the intervention ^14,41,64^.

We conducted a meta-analysis to evaluate the effects of MI on emotional states in individuals with BD, synthesizing data across 12 emotional outcome domains, including depression, anxiety, emotional well-being, and self-care (**Figure 6**). The pooled analysis revealed a small but statistically significant overall effect favoring the intervention group (MD = 0.20; 95%CI: 0.04−0.36), with a z-score of 2.46 and a *P*-value of 0.01. Moderate heterogeneity was observed (I^2^ = 49%, τ^2^ = 0, *P*-value = 0.02), indicating variability in effect sizes across studies. The test for subgroup differences across outcome domains (e.g., depression, pain, anxiety, emotional well-being, and self-care) was not statistically significant (χ^2^ = 14.47, df = 11, *P*-value = 0.21), suggesting that no single domain demonstrated a consistently stronger effect than others.

Within the depression-related outcomes, results were mixed. The PHQ-9 and BDI-II indicated positive effects, whereas the CES-D yielded a small negative effect (MD = -1.10; 95%CI: -5.58−3.38), and the 95%CIs for anxiety burden and pain included zero, indicating non-significance.

For emotional well-being, the study by Palumbo et al. ^52^ showed a meaningful and statistically significant improvement (MD = 3.44, 95%CI: 0.19–6.69). Stroke Impact Scale Emotions (SIS-Emotions) scores also showed a positive pooled effect (MD = 4.82; 95%CI: -2.48−12.11), although with high heterogeneity (I^2^ = 78%, τ^2^ = 31.34, *P*-value = 0.01) and a non-significant test for subgroup effect (z-score = 1.29, *P*-value = 0.20). Self-care domains, assessed through the Strategies Used by People to Promote Health (SUPPH) subscales, consistently showed positive mean differences, but most had wide confidence intervals that crossed zero, indicating statistical non-significance.

Despite variability across individual outcomes, the results support the potential of MI to enhance emotional well-being and related domains in individuals with BD. However, the presence of moderate to high heterogeneity in several subgroups highlights the need for cautious interpretation and suggests that further research is needed to clarify which domains and populations benefit most.

*2.5. Quality of Life*

Notably, some studies identified enhanced QoL as a direct outcome of MI ^5,14,21,23,42,51,52^, including many global outcomes such as psychological wellbeing, physical recovery, or social engagement.

We carried out a meta-analysis to evaluate the impact of MI on QoL in individuals with BD, drawing on data from five studies and three standardized QoL measures (**Figure 7**): the Stroke-Specific Quality of Life Scale (SS-QoL), the Quality of Life after Brain Injury (QoLIBRI), and the Quality-of-Life Index (QoI). The pooled findings offer a cautiously optimistic view, though overall results did not reach statistical significance (z-score = 1.45; *P*-value = 0.15). The overall effect size favored the music intervention group, with a pooled mean difference of 4.44 (95%CI: -1.56–10.43). The overall heterogeneity across studies was substantial (I^2^ = 73%, τ^2^ = 34.06, *P*-value < 0.01), indicating variability in effect estimates. The test for subgroup differences between the three QoL measures (SS-QoL, QoLIBRI, and QLI) was not significant (χ^2^ = 1.56, df = 2, *P*-value = 0.46), suggesting that neither measure consistently outperformed the other in capturing treatment effects. In the SS-QoL subgroup, three studies were included. Cha et al. ^5^ reported a large and significant benefit (MD = 18.90, 95%CI: 8.40–29.40), while Jeong (2007) reported a negligible and non-significant effect (MD = -0.05, 95%CI: -0.46–0.36), and Grau-Sánchez (2018) indicated a moderate but non-significant improvement (MD = 7.40, 95%CI: -5.62–20.42). The pooled estimate from this subgroup yielded a mean difference of 7.86 (95%CI: -3.66–19.38), indicating a trend toward benefit but lacking statistical significance (z-score = 1.34, *P*-value = 0.18). Notably, heterogeneity in this subgroup was substantial (I^2^ = 85%, τ^2^ = 82.72, *P*-value < 0.01), suggesting that differences in study design or sample characteristics may explain the variation in observed effects. For the QoLIBRI subgroup, the analysis included a single study ^48^, which reported a small, non-significant improvement in QoL (MD = 2.80, 95%CI: -4.33–9.93). Finally, the QLI subgroup, represented by Palumbo et al. ^52^, also found a minimal and non-significant effect (MD = 0.74, 95%CI: −2.10–3.58). The lack of additional studies in these subgroups limits broader generalization, but it contributes to the overall model assessing QoL effects across both tools. Overall, the meta-analysis suggests a potential benefit of MI on QoL in individuals with BD, though current evidence remains inconclusive due to limited studies and high heterogeneity.

*2.6. Global improvement*

The outcome of global improvement has recently been considered an important variable in studies investigating music-based rehabilitation for ABI ^11,50,52^. However, previous reviews did not consider it as a measure of global improvement ^65^. The variability in the concept of global improvement is evident, as some measures focus on comprehensive cognitive assessments, such as the Clinical Dementia Rating (CDR) or the Global Deterioration Scale (GDS) ^50^, while others assess the level of disability in daily activities, such as the Functional Independence Measure (FIM) ^11^. Palumbo et al. ^52^ evaluated global post-stroke disability through clinical interviews using the modified Rankin Scale (mRS). However, the SIS, a self-reported instrument, is widely recognized for complementing the understanding of overall recovery and global improvement ^25,52^.

The meta-analysis on global improvement outcomes (**Figure 8**) across four studies found no statistically significant overall effect of MI in individuals with acquired brain injury (MD = 0.03, 95%CI: −0.34-0.41, z-score = 0.18, *P*-value = 0.86), with moderate but non-significant heterogeneity (I^2^ = 31%, *P*-value = 0.20). Subgroup analysis by outcome measure (CDR-SB, GDS, FIM, mRS, and SIS) revealed no significant differences (χ^2^ = 6.61, df = 4, *P*-value = 0.16), though the SIS Recovery subgroup showed a positive trend with marginal statistical significance (z-score = 1.81, *P*-value = 0.07) and no heterogeneity (I^2^ = 0%). Current evidence does not support a significant global improvement effect from MI on AB populations across the included measures. However, the SIS Recovery domain showed a promising trend that warrants further investigation, particularly in larger, more targeted studies.

3. Types of musical interventions in brain damage studies

This review examines the types of musical stimulation used in music-based therapies for acquired brain injury (ABI), most of which focus on rehabilitation, with the goal of linking specific interventions to reported outcomes across 70 studies (**Section 3.2**). A major challenge in assessing the efficacy of MI lies in the substantial variability in their duration, ranging from under an hour to, in rare cases, over 60 hours (**Table 1**). Notably, only seven studies implemented long-term interventions (≥30 hours), and these were consistently associated with significant improvements across multiple domains. These included: (i) MST ^20,42^, *ii*) Rhythm- and music-based therapy ^13^, *iii*) Choral and singing therapy ^54^, and *iv*) Music listening interventions ^26,33^. Active music interventions, where participants engage directly with music through instrument playing or singing, were the most prevalent among these high-impact studies (**Table 1**).

*3.1. Interventions involving musical instrument playing*

In 31 studies, active music training, such as playing a musical instrument, was a fundamental component of restorative therapy targeting both fine and gross motor upper-limb skills. Among these, 21 studies reported significant improvements in UEF in individuals with BD. Piano playing, in particular, has been highlighted as an effective intervention for hand rehabilitation ^8,9,11,12^ as well as for cognitive enhancement in executive functions such as attention, learning strategies, and memory retrieval ^41,66^. Grau-Sánchez et al. ^67^ reported notable improvements in keyboard task performance after just one session of piano playing, while Segura et al. ^25^ confirmed enhanced motor recovery through enriched MST compared to a conventional motor program.

Moreover, 24 studies implemented MI techniques involving instrumental playing. Among these, MST, which typically includes playing electronic instruments such as a MIDI keyboard and a drum set, was the most frequently applied approach, reported in 14 studies ^10,16,19-24,42,45,51,67-72^. Additionally, NMT demonstrated the rehabilitative potential of active MI in seven studies ^45,46,73^.

*3.2. Interventions combining music and movement*

Restoring walking ability is a key milestone in cerebral damage rehabilitation, and MI involving movement synchronized with music has demonstrated efficacy in motor recovery. Seven studies focusing on rhythmic stimulation reported significant gait improvements ^2-6,13^. Among these, RAS and Fast Tempo Auditory Stimulation (FAST), both of which typically incorporate a metronome and sometimes preferred music, were the most used techniques. Also, five studies reported an improvement in UEF thanks to interventions based on music and movement. A novel music-based rehabilitation approach, musical sonification of arm movements, has also been explored. However, results remain preliminary, with only limited evidence supporting its effectiveness in enhancing movement smoothness of arms and hand function ^15,17,18^.

*3.3. Interventions based on singing*

Singing therapy has played a prominent role in 11 studies, with interventions including: *i*) Choir participation ^54,56^, *ii*) Melodic Intonation Therapy (MIT) exercises ^28,29,31,32,35,36^, *iii*) Singing favorite songs with guitar accompaniment ^34,57^, and *iv*) Intensive voice training, incorporating intonation, prosody, breathing, rhythm, and improvisation ^30^. The primary benefit of singing-based interventions is enhanced communication abilities in aphasic patients, with multiple studies demonstrating that singing is more effective than conventional language rehabilitation therapies ^28-32^.

*3.4. Interventions involving passive music listening*

Music listening, as a passive intervention in which participants are exposed to recorded music, has been examined in 12 experimental studies employing various musical genres and styles, including: *i*) popular songs ^39,59^, *ii*) favorite music of participants ^26,27,40^, and *iii*) comparisons between different types of music ^55,58,62^, such as vocal *vs.* instrumental ^33^ or classical *vs.* preferred music ^55^ and *iv*) relaxing music ^60,62^. The main benefits of music listening were emotional well-being, reported in seven studies, and cognitive function, reported in six studies (**Table 1**), with five of those focusing on memory enhancement. Sihvonen et al. ^33^ suggested that listening to vocal music is an effective, easily applicable tool for language and cognitive recovery after a stroke. Additionally, communication and behavioral improvements were reported in five studies, further demonstrating the diverse benefits of this low-demand intervention.

60.	Palmquist ET, Underbjerg M, Ridder HM, Jespersen KV. Music listening for improvement of sleep in post-acute rehabilitation of adults with acquired brain injury: A feasibility study

. *NJMT*. 2025;34(3):245-260.
